# Machine Learning for Antibiotic Stewardship in the Treatment of Stapholycoccus Bacterial Infections

**DOI:** 10.1101/2022.11.28.22282797

**Authors:** Trevor J. Brokowski, Jeffrey N. Chiang

## Abstract

Antibiotic resistance is one of the leading issues in modern healthcare due to the inability to treat common infections with available antibiotics. Many of the mechanisms of resistance have been caused by the inappropriate prescription of antibiotics to treat illnesses such as the cold or flu or the over-prescription of broad-spectrum antibiotics. Epitomizing this problem is the Staphylococcus bacteria where certain strains have become resistant to penicillin-related drugs and Vancomycin, one of the treatments for MRSA. To address this, we developed machine learning models to predict antibiotic activity and susceptibility using a patient’s entire available electronic health record. We selected patients who were suspected of having a staph infection from the Medical Information Mart for Intensive Care III (MIMIC-III) data set and utilized their microbiological culture results to identify the number of patients that were prescribed an inappropriate antibiotic and then propose suitable alternatives. In our test set, we identified that empiric prescriptions had an efficiency rate of 40 percent (the rate at which an antibiotic that would provide activity was prescribed), and the other 60 percent of cases were not susceptible to the prescribed antibiotic or the antibiotic that they were given was not tested for susceptibility against their infection. Our best models identified antibiotic susceptibility with AUROCs up to 0.9 and raw specificity up to 0.7. The models were also able to propose suitable alternatives in all but 10 cases. Overall these results demonstrate the need for implementing clinical decision support systems advising clinicians during the prescription process, and our further work will address this issue.

## 1. Introduction

The emergence and spread of drug-resistant pathogens threaten global health by affecting our ability to treat common infections. ^1^ Although genetic changes can induce the development of antimicrobial resistance over time, the rapid development of the mechanisms of resistance can be attributed to the inappropriate use of common and broad-spectrum antibiotics.^2–5^ For decades, doctors have prescribed antibiotics to treat colds, the flu, and other viral infections that don’t respond to antibiotics, and even when used properly, many organisms are capable of rapid evolution to gain resistance to these drugs. Despite past achievements in minimizing the development and spread of drug-resistant organisms, the inappropriate over-prescription of antibiotics during the coronavirus 2019 pandemic (COVID-19) has resulted in a significant increase in hospitalizations due to multi-drug-resistant organisms, and, by 2050, the World Health Organization (WHO) estimates that antibiotic-resistant infections will cause of more than 10 million deaths per year.^6–7^

To combat the spread of resistance, the Centers for Disease Control and Prevention (CDC) have stated that the most important action is improving antibiotic prescription through antibiotic stewardship. ^8^ Common strategies involve reducing the amount and the duration of unnecessary antibiotic treatment. Yet, despite these initiatives, it is estimated that up to 60 percent of clinicians still prescribe antibiotics in situations where their use is either unnecessary, the incorrect antibiotic was prescribed, or the antibiotic was administered at the incorrect dose or for the incorrect amount of time.^9–10^

There is also growing concern over a certain class of antibiotic-resistant infections known as “Superbugs”. These multi and pan-resistant bacteria no longer respond to treatment with available antibiotics and the looming threat is more bacterial organisms will follow.^11^ One of the “Super-bugs”, methicillin-resistant Staphylococcus aureus (MRSA), is a strain of Staphylococcus aureus (S. aureus) that has developed resistance to many of the antibiotics frequently used to treat staph infections. In the last year, MRSA directly caused the death of over 100,000 people and is gaining resistance to other forms of treatment.^12,13,14^ If this phenomenon persists, routine medical practices and procedures will be crippled, drastically increasing infection-related mortality and treatment costs.

Clinical decision support systems present ample opportunity to help clinicians adhere to antibiotic stewardship measures in practice.^15^ Historically, these systems have not been adopted due to the difficulty of integration into clinical workflows and their inability to adapt to dynamic local antibiotic resistance patterns.^16^ However, modern hospital IT infrastructures and electronic health record software (EHR) make it possible to integrate clinical decision support systems and utilize continuously integrated and continuously deployed machine learning models for clinical decision-making. With these advancements, it is possible to implement clinical decision-support systems to ensure that clinicians are aware of antibiotic stewardship measures when prescribing antibiotics to treat staph and other bacterial infections.^17^

Electronic Health Records present the means by which we can survey past events in order to evaluate the quality of care and help improve future practices. There have been extensive studies using EHR data to deliver precision care through clinical decision support systems, and, among these, many have focused on utilizing microbial culture results found in electronic health records to improve antibiotic stewardship measures in clinical settings.^18,19,20^

Here, we seek to investigate the use of machine learning-driven approaches for predicting antibiotic susceptibility to aid in empiric antibiotic therapy to one of the most prevalent drug-resistant organisms in hospitals around the world. Specifically, we seek to predict whether an antibiotic will provide activity against the bacterial staph infection, evaluate hospital performance in prescribing appropriate and effective antibiotics to treat the infection, and then identify relevant predictors of activity in the tested antibiotics. Using Electronic health record data, we can tailor antibiotic therapy toward individual patients to prescribe the most effective and specific antibiotics. We hope that this method will result in an actionable solution for promoting antibiotic stewardship among patients admitted with staph infections to improve treatment and reduce the opportunity for more resistant strains to develop.

## 2. Materials and Methods

### 2.1 Data Sources

The data used in this analysis was extracted from the Medical Information Mart for Intensive Care III (MIMIC-III). The MIMIC-III database contains deidentified health-related data associated with over forty thousand patients who were admitted to the critical care units of the Beth Israel Deaconess Medical Center between 2001 and 2012. The data mart was deidentified and structured in accordance with Health Insurance Portability and Accountability Act (HIPAA) standards containing patient demographics, comorbidities, procedures, medications, laboratory test results, vital signs, microbiology data, discharge summaries, and International Classification of Disease, 9th Edition (ICD-9) codes. ^21^

### 2.2 Cohort Definition

Patients who were assumed to have staph infections met the inclusion criteria for this study. The exact phenotype required for inclusion in the study was any microbiological culture that returned positive for a staph-related organism obtained from any of the following: blood, urine, cerebral spinal fluid, pleural cavity, or joint, and the prescription of an antibiotic that was subsequently tested for susceptibility. Staph infections, including Staph aureus positive, Staphylococcus Coagulase-negative, and MRSA present identifiable phenotypes such as painful red bumps on the skin and feverish conditions - among many others.^22,23^Due to these visible and identifiable phenotypes, we make the assumption that the clinician has a high suspicion that the identity of the bacterial infection is staph related, the microbiology test will return positive for a staph-related organism, and empiric therapy will be oriented towards staph-related treatments. Moreover, patients with multiple ICU admissions that met the study criteria were analyzed independently and assigned to the same train/test split to prevent test set contamination. Figures 2 and 1 illustrates the flow chart of the data collection and the prediction timeline.

**Figure 1.**
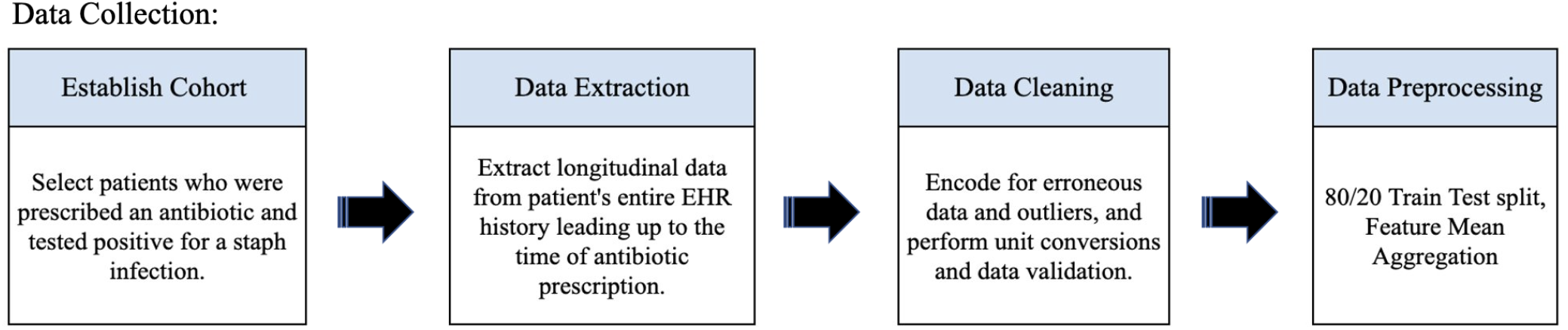
Data Collection Overview: Raw Data is taken from the MIMIC-III database. A cohort consisting of adult patients who tested positive for a staph-related infection and were prescribed an antibiotic was selected. Longitudinal data were then extracted from their entire electronic health record, and then cleaned and pre-processed for model training and testing.

**Figure 2.**
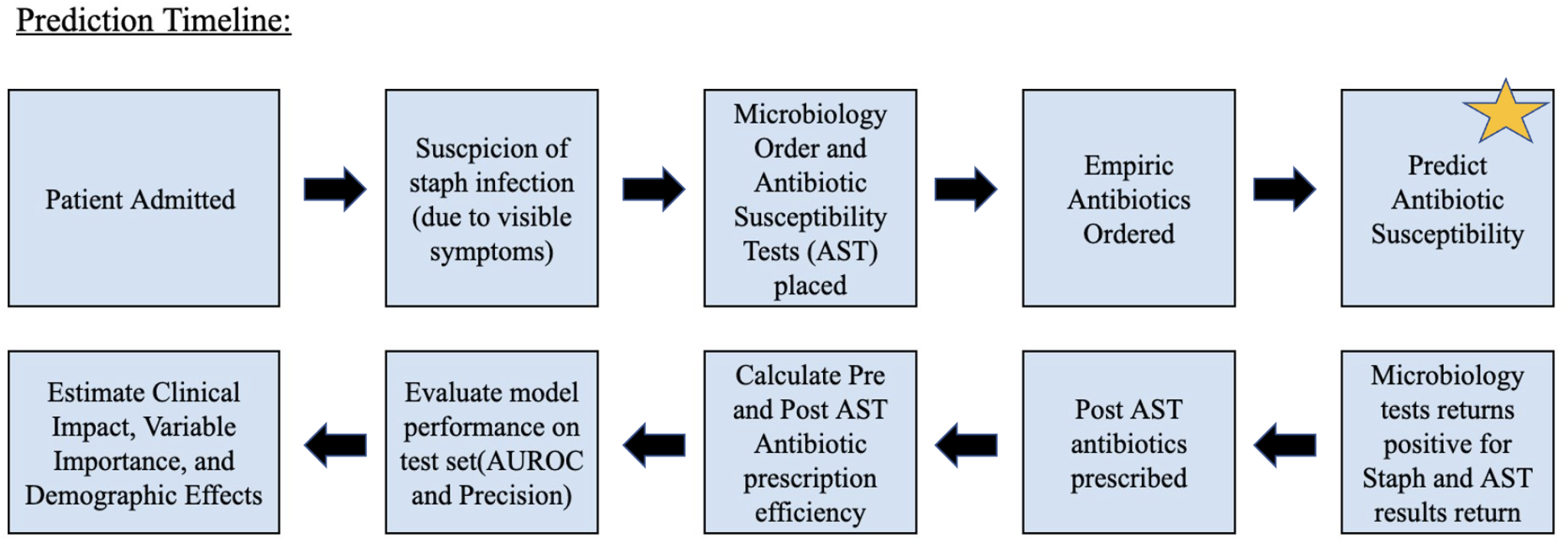
Prediction Timeline: Here we present the proposed prediction timeline for the classifier. First, a patient is admitted into the ICU and then the patient develops symptoms of a staph infection. Microbiology and AST cultures are ordered and then empiric prescriptions are given. Upon the time of antibiotic prescription, we make predictions on antibiotic susceptibility to guide empiric antibiotic regimens. Then we evaluate the clinician efficiency of prescription in both empiric and specific antibiotic regimens.

**Figure 3.**
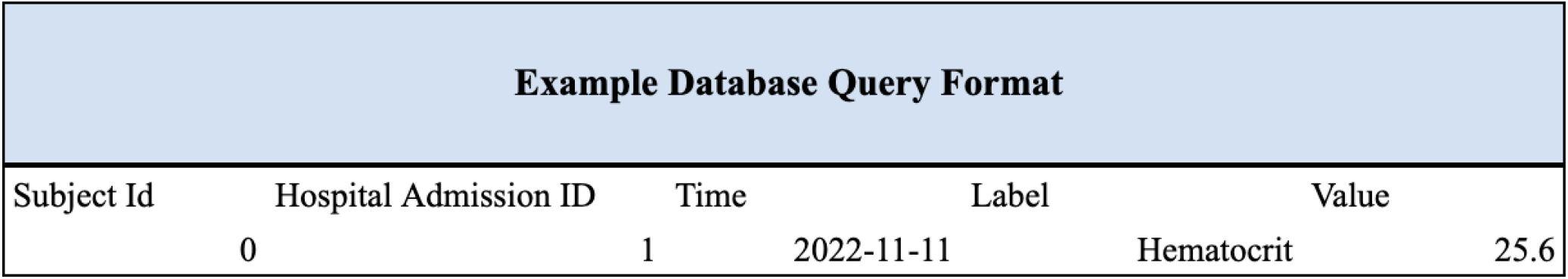
The longitudinal data is extracted such that we create a table consisting of the subject and hospital admission identifiers, the time of data collection, and then the type of data (corresponding to label) and the value of that label. This allows us to select all relevant patient information by specifying the patient, the time, and the type of information.

### 2.3 Feature Engineering

We developed a single data structure that was used for all of the predictions instead of manually curating datasets for each susceptibility profile. The data was extracted in a long format and organized by the respective table, patient id, and time of data. Namely, we extract the subject id, the hospital admission id, the time of the event, the type of event, and the value of the event. An example from the chart events table is presented in 3.

The raw clinical data was then cleaned and formatted to address data quality issues such as erroneous values and the disarray of varying units. For example, the units of the amount of a type of drug given were all converted to mg, and erroneous values such as “error” in the chart events table were encoded as -1 and considered to be missing. We then construct a feature matrix with the available static and longitudinal data using the extracted and cleaned clinical information. Static data includes features such as gender, ethnicity, language, insurance, and religion. Longitudinal data includes features from the lab events, chart events, input events (medication), prescriptions (medication), diagnoses, procedures, and microbiology events tables in the MIMIC-III database. The data was then encoded to address the numeric and categorical features. Categorical information included ICD 9 codes extracted from the diagnoses and procedures table, as well as some of the features in the chart events and lab events tables. These were then encoded to dummy variables using the one-hot-encoding technique. Continuous features included values from the input events, prescriptions, and chart events tables. If a feature was not present for a patient, the value was encoded as -1 and handled the same way as erroneous data. This allowed us to encode missing values into our data without needing to utilize data imputation methods.

The value -1 was chosen instead of 0 because some of the observations possessed the measured value 0. To construct the final data frames, we used all of the available patient data before the time of the prescription and then aggregated the values by taking the non-filler (−999) mean of each of the features. In total, there were 4,754 features included in the final dataset.

### 2.4 Labeling

We trained 9 binary machine learning models to estimate the probability that an antibiotic selection would provide activity against the patient’s infection, namely one model per antibiotic. We encode our positive class as the ability of the antibiotic to provide activity against the infection and encode our negative class as the inability of the antibiotic to provide activity against the bacterial infection (if the AST returned intermediate or resistant). If the antibiotic was not tested against the patient’s infection we assume that the clinician would not consider prescribing that antibiotic and so we also encode this into our negative class.

The unit of observation in our study is the start of a unique antibiotic regimen for the patients in our cohort, and we mark the prediction time to be the first time at which a unique empiric antibiotic (antibiotics prescribed before AST results were available) was entered into the patient’s electronic health record signifying the start of a treatment regimen. We then retrospectively evaluate the specific antibiotic regimens (antibiotics prescribed after AST results were available). The purpose of this is to evaluate hospital performance in prescribing appropriate antibiotic regimens and to evaluate the allocation of antibiotics in empiric and post-AST treatment regimens.

An antibiotic regimen was labeled appropriate if it would provide activity against the bacterial infection, and an antibiotic regimen was said to provide activity if all microbial organisms that grew in the patient’s culture were susceptible to the antibiotic. Activity is measured in terms of the minimum inhibitory concentrations evaluated in the antibiotic susceptibility testing procedure. Similarly, an antibiotic regimen was labeled inappropriate if the microbiological culture was labeled as resistant or intermediate to the prescribed antibiotic or if the prescribed antibiotic was not tested against the patient’s infection. We define efficiency to be the fraction of appropriate antibiotic prescriptions to the total number of prescriptions. As our unit of observation is the susceptibility profile of the antibiotic, we are constrained by the number of times that an antibiotic was included in a susceptibility test. So, due to the fact that not all antibiotics that were prescribed were tested for susceptibility, we are limited in the number of antibiotics for which we can estimate the probability of activity against the infection. Thus, we only include antibiotics that were prescribed to the patients in the cohort and were subsequently tested and found to provide activity against more than 2.5 percent of the cohort’s infections. These antibiotics include Levofloxacin, Clindamycin, Vancomycin, Linezolid, Oxacillin, Gentamicin, Erythromycin, and Daptomycin.

### 2.5 Training and Model Selection

The patients that were prescribed an antibiotic before the results from an antibiotic susceptibility test were included in the model training and testing procedure. This group was then split by unique hospital admission identifiers using an 80/20 train test split. This was done to avoid test set contamination where we would include the same patient in the train and test set if they had been issued two distinct antibiotics before their AST results had returned. We selected two tree-based models to perform the analysis of the data set: random forest and gradient-boosting trees. This was done because prior results have demonstrated that tree-based methods often outperform linear and logistic regression models due to their ability to model non-linear interactions with high-dimensional data.^18,19^The random forest models were fit using the sci-kit learn python package and the gradient-boosted tree models were fit using the lightgbm python package. ^24,25^ The model training procedure utilized a grid search over the training set to identify the hyperparameters that led to the highest mean area under the receiver operating characteristic curve (AUROC). The binary cross entropy loss function was used to evaluate the predictions, and the final model was chosen by selecting the model with the highest AUROC on the test set.

## 3. Results

### 3.1 Cohort

Our selected cohort contains N = 7655 prescriptions from 4734 unique ICU admissions in the MIMIC-III data that met the inclusion criteria for the study. Of the original N prescription regimes, 5185 (68 percent) were made empirically to 3795 patients before AST test results were available, and 2470 (32 percent) prescriptions were made to 1968 patients after AST results were available. There were 1047 patients who were prescribed an antibiotic before and after their AST results were available. The train/test split based on the ICU admission id resulted in a train set containing N = 3635 prescriptions to 2656 patients and a test set containing N = 1550 prescriptions and 1139 patients. Table 2 summarizes the demographic breakdown of these patients across the train/test split.

### 3.2 Model Performance

In 4, we report the performance of the antibiotic susceptibility classifiers across the test set and show the prevalence (the fraction of patient infections for which the antibiotic was listed as susceptible across the test set), the average precision, and the area under the receiver operating characteristics (AUROC) for the 9 antibiotics. The best model refers to the model that achieved the highest AUROC across the test set.

**Figure 4.**
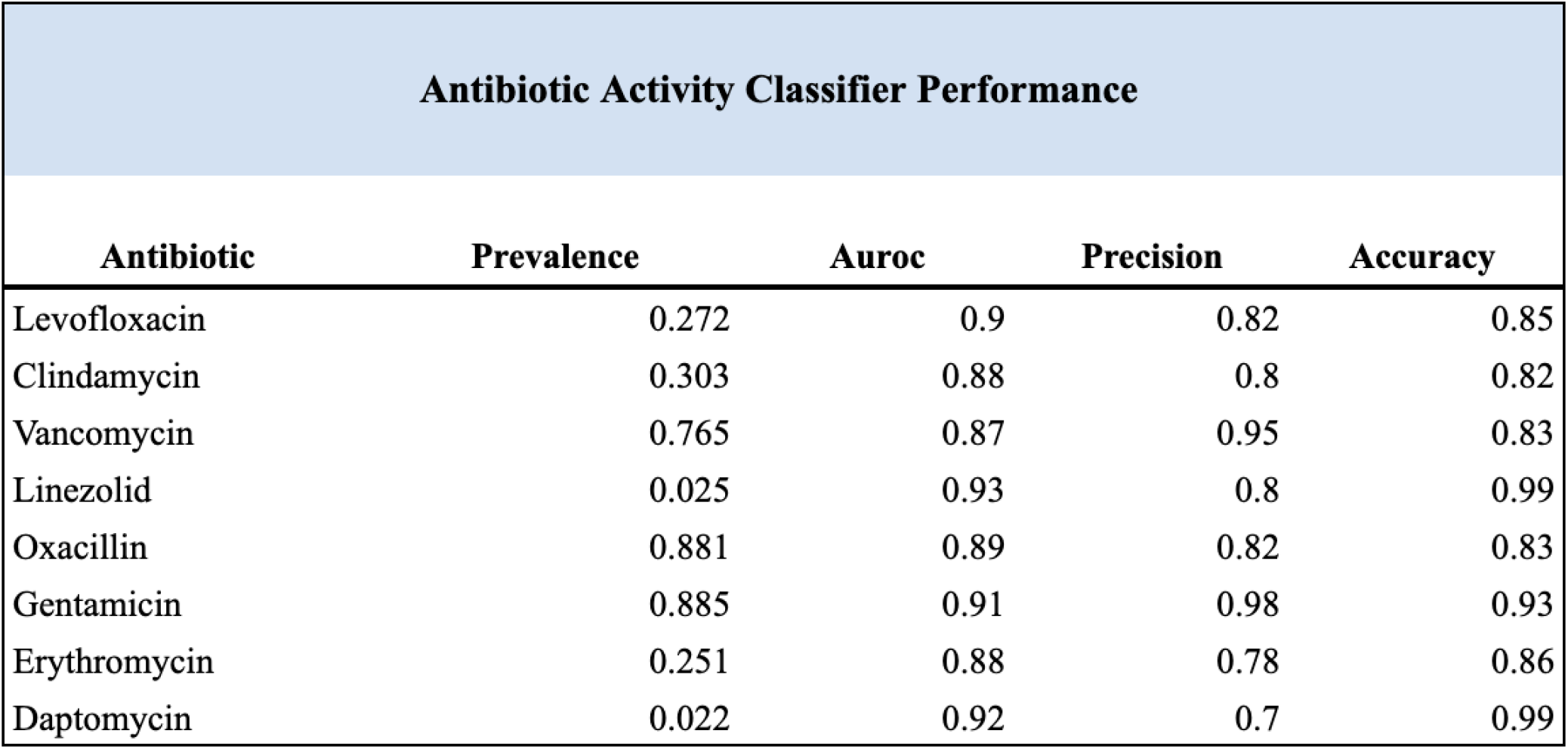
Here we report the prevalence (the total amount of susceptible antibiotics from the cohort’s cumulative antibiograms), Area Under the Receiver Operator Characteristic Curve, Precision, and Accuracy for each of the 9 binary antibiotic susceptibility classifier models.

**Figure 5.**
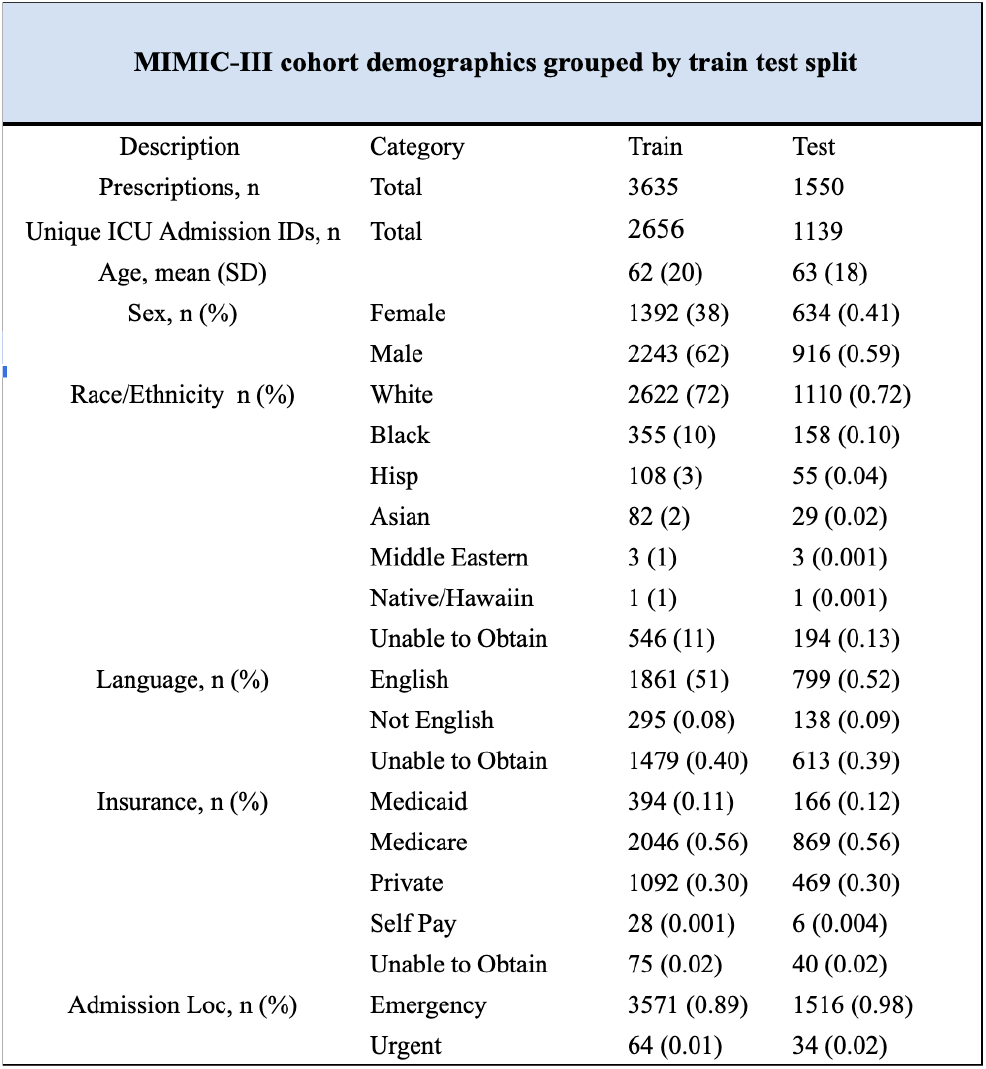
The demographic information corresponding to the 80/20 train and test set for our cohort

The Gradient Boosting Tree slightly outperformed the random forest classifier across all of the classifiers where the average precision ranged from 0.55 to 0.98 and the AUROC ranged from 0.88 to 0.98. In 6, we display the ROC curve and the Precision-Recall Curve from the Vancomycin Susceptibility Estimator. We then retroactively compute high specificity thresholds of 0.95 for each classifier and apply them to the test set in order to predict the antibiotics that would provide activity for each patient’s infection.

### 3.3 Clinical Evaluation and Model Utility

To evaluate the potential utility of our model in the clinical setting we first sought to evaluate the rate at which clinicians prescribed appropriate antibiotics - antibiotics that would provide activity against the staph infection - as well as evaluate the allocation of the different antibiotics included in the study. We retroactively compute the fraction of patients that were given antibiotics that would provide activity for their bacterial infection in both our train and test set. We then examine the allocation of the prescribed antibiotics to evaluate the hypothesis that empiric antibiotic therapy for patients with symptoms of a staph infection will be prescribed antibiotics relevant to the treatment of staph. Then, for our test set, we evaluate the number of available antibiotics that our model predicted would provide activity against bacterial infection using the high specificity threshold. 7 and 8 present the results of the empiric antibiotic therapy issued to the patient. We define empiric therapy as having an antibiotic regimen started before the results from an antibiotic susceptibility test had returned - the time stamp for the first issued antibiotic being before the return of the antibiotic susceptibility test. These results verify our hypothesis that empiric antibiotic therapy for the treatment of staph infections will be biased toward drugs known to provide activity against staph infections - namely Vancomycin and Levofloxacin which constitute nearly 80 percent of the total prescribed antibiotics for the cohort. We also report the efficiency of the clinician performance which we define as the fraction of appropriate prescriptions from the total amount of prescriptions per drug. We see that on average, between the training and the test set, clinician performance has an average of approximately 41 percent efficiency and prescribe antibiotics that will provide activity against the bacterial infection less than half the time.

**Figure 6.**
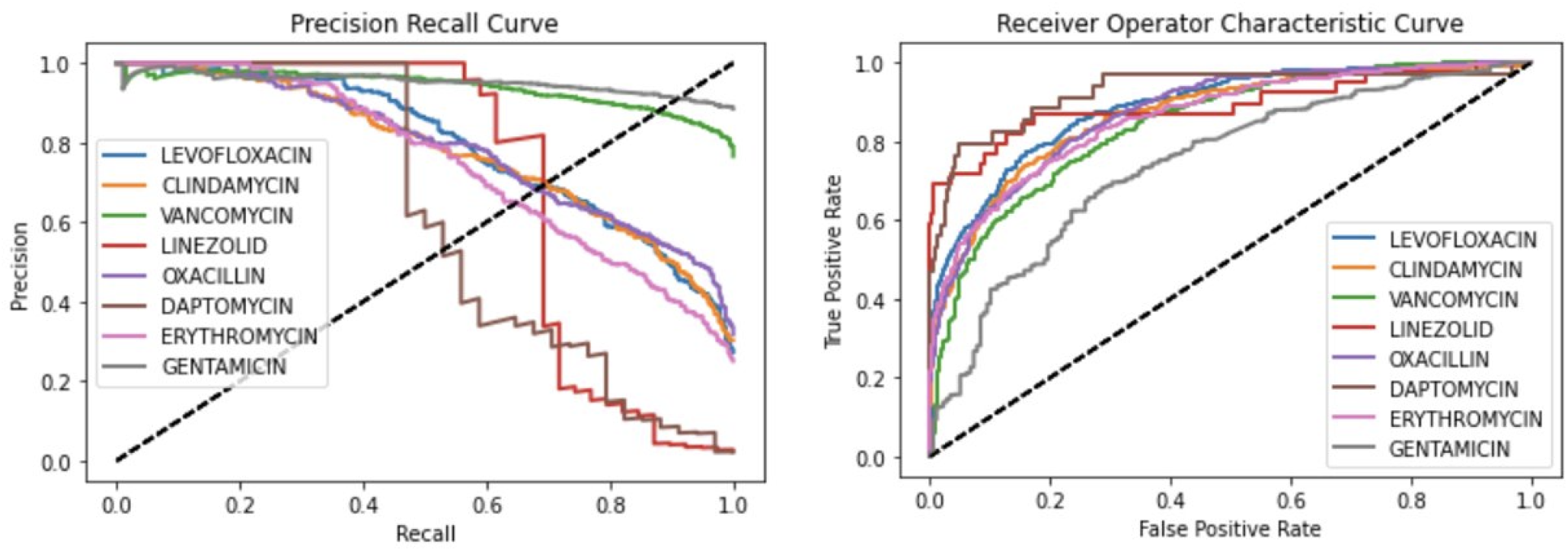
Receiver operating characteristic curves and Precision-recall curves for all models. We use different colors to differentiate the models.

**Figure 7.**
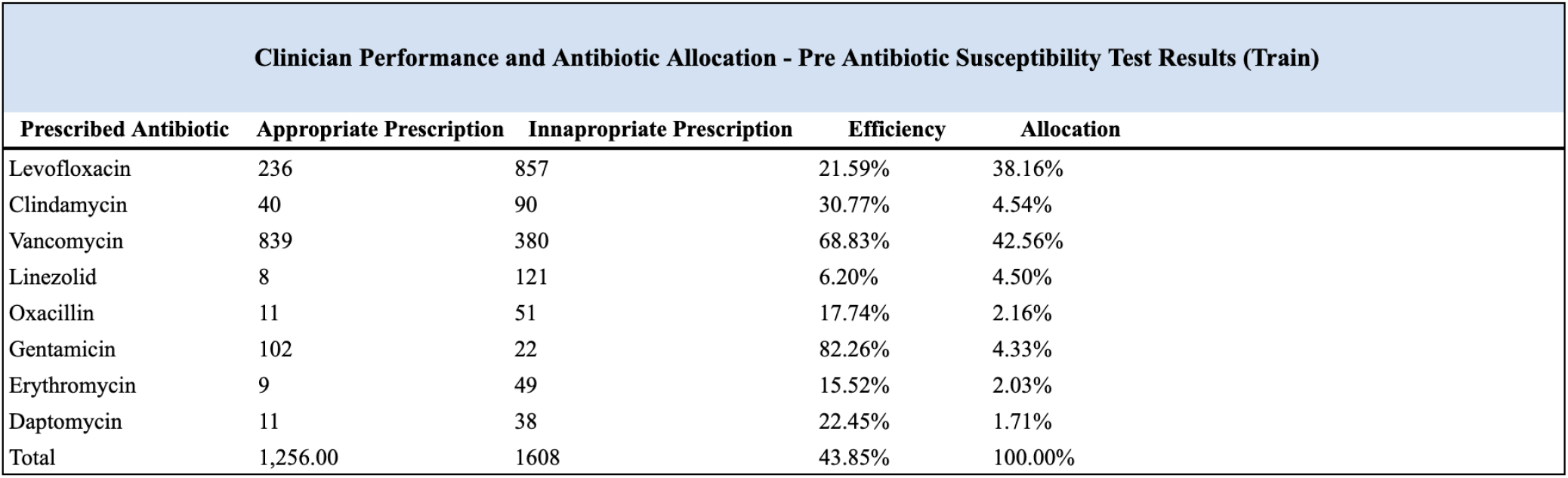
We break down the relative antibiotic allocation, the number of appropriate prescriptions (patient was susceptible towards the prescribed antibiotic), inappropriate prescriptions, and the efficiency rate of prescriptions for each antibiotic used in the study. These are the results across the train set which provide a larger data set

Our model is able to estimate the probability that the antibiotic will provide activity against the patient’s staph infection. In table 6, we report the median of the predicted number of drugs that could have provided activity when an inappropriate antibiotic was prescribed. In our test set, we found there to be 751 total cases of inappropriate empiric antibiotic prescriptions (59 percent of the total prescriptions), and our model predicted that an average of 3.17 antibiotics could have provided activity against the infection, compared to the true average of 3.51 antibiotics. Moreover, there were only 10 predicted events in which no antibiotic would provide activity against the bacterial infection. Using our high specificity threshold, we expect that this model could have theoretically benefited close to 60 percent of patients. In 9, we conducted a retrospective analysis of clinician performance in efficient antibiotic prescription and allocation after clinicians had received the antibiotic susceptibility test results. This was done in order to examine the rate at which clinicians adhere to antibiotic stewardship guidelines as well as evaluate the potential impact of making clinicians aware of antibiotic susceptibility tests each time at which a prescription regimen was started. We find that prescription efficiency does not increase across the board. We find notable improvements in the prescription of Oxacillin and Clindamycin, and efficiency decreases in Vancomycin. We do see a slightly better diversification of the prescribed antibiotics with Vancomycin and Levofloxacin constituting 60 percent of the total number of regimens. Similarly, we calculate the number of appropriate antibiotics that could have been prescribed in place of an inappropriate prescription using the returned antibiotic susceptibility tests. Similar to the predicted results in the empiric test set we find that we could have prescribed a median of 3 antibiotics that would have provided activity instead of the inappropriately prescribed antibiotic.

**Figure 8.**
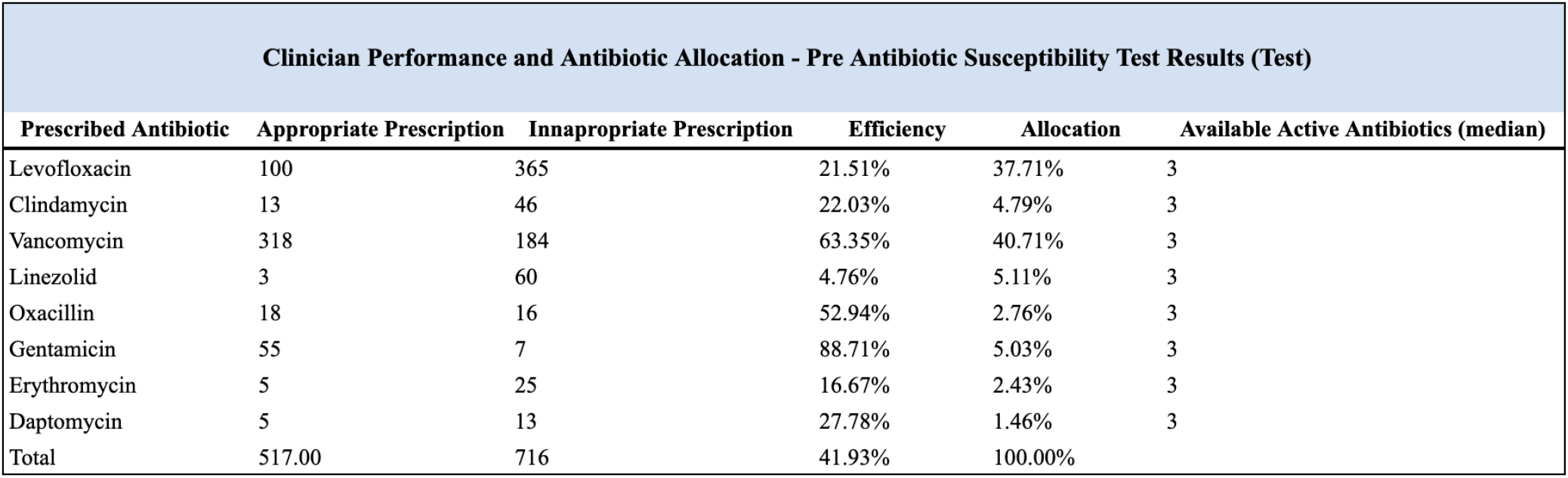
We break down the relative antibiotic allocation, the number of appropriate prescriptions (patient was susceptible towards the prescribed antibiotic), inappropriate prescriptions, and the efficiency rate of prescriptions for each antibiotic used in the study. These are the results across the test set. At each time the antibiotic was prescribed, the available active antibiotic column sums the total number of antibiotics that all of the classifiers predicted would provide activity. Thus, we can view this column as suitable alternatives in cases where an inappropriate antibiotic was prescribed.

**Figure 9.**
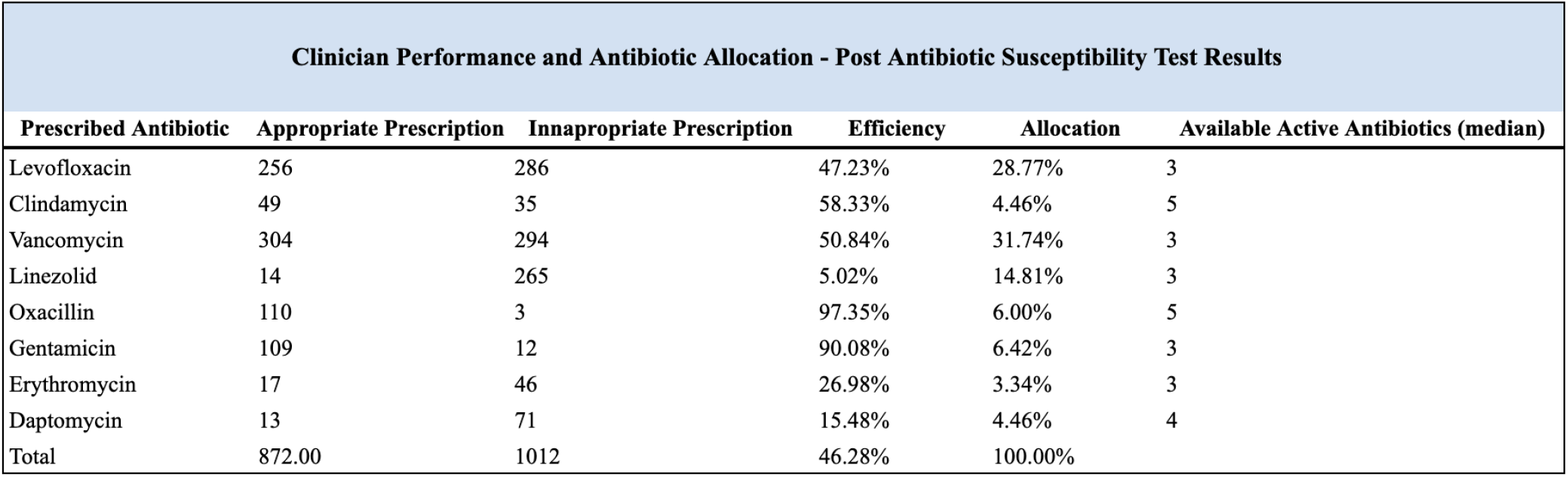
We break down the relative antibiotic allocation, the number of appropriate prescriptions (patient was susceptible towards the prescribed antibiotic), inappropriate prescriptions, and the efficiency rate of prescriptions for each antibiotic used in the study. These are the results found from specific antibiotic treatment regimens (post-AST). At each time the antibiotic was prescribed, the available active antibiotic column sums the total number of antibiotics that would have provided activity found from the antibiotic susceptibility tests. Thus, we can view this column as suitable alternatives in cases where an inappropriate antibiotic was prescribed.

After examining the potential room for improvement of prescription efficiency using our model, we then examined the features that were most influential in the classification process. We calculate Shapley values, the average marginal contribution of a feature value across all possible coalitions.^26^This allows us to examine the feature importance in high-dimensional and non-linear spaces. We calculate these values for each of our antibiotic classifiers and then select the most impactful features across each classifier. 10 presents our results. Through these interactions, we are able to discern that the three categories contributing to susceptibility are the prior antibiotics prescribed to a patient, blood test-related features, the presence of a prior antimicrobial-resistant infection (V09), and the age demographics and the ability of the patient to speak English.

Finally, we evaluate the effect of demographic information on the classifier with results in 11. We find notable decreases in the AUROC of the classifiers when the demographic information is removed suggesting that factors such as race, language, and insurance play a role in predicting the activity of an antibiotic for a patient’s infection.

**Figure 10.**
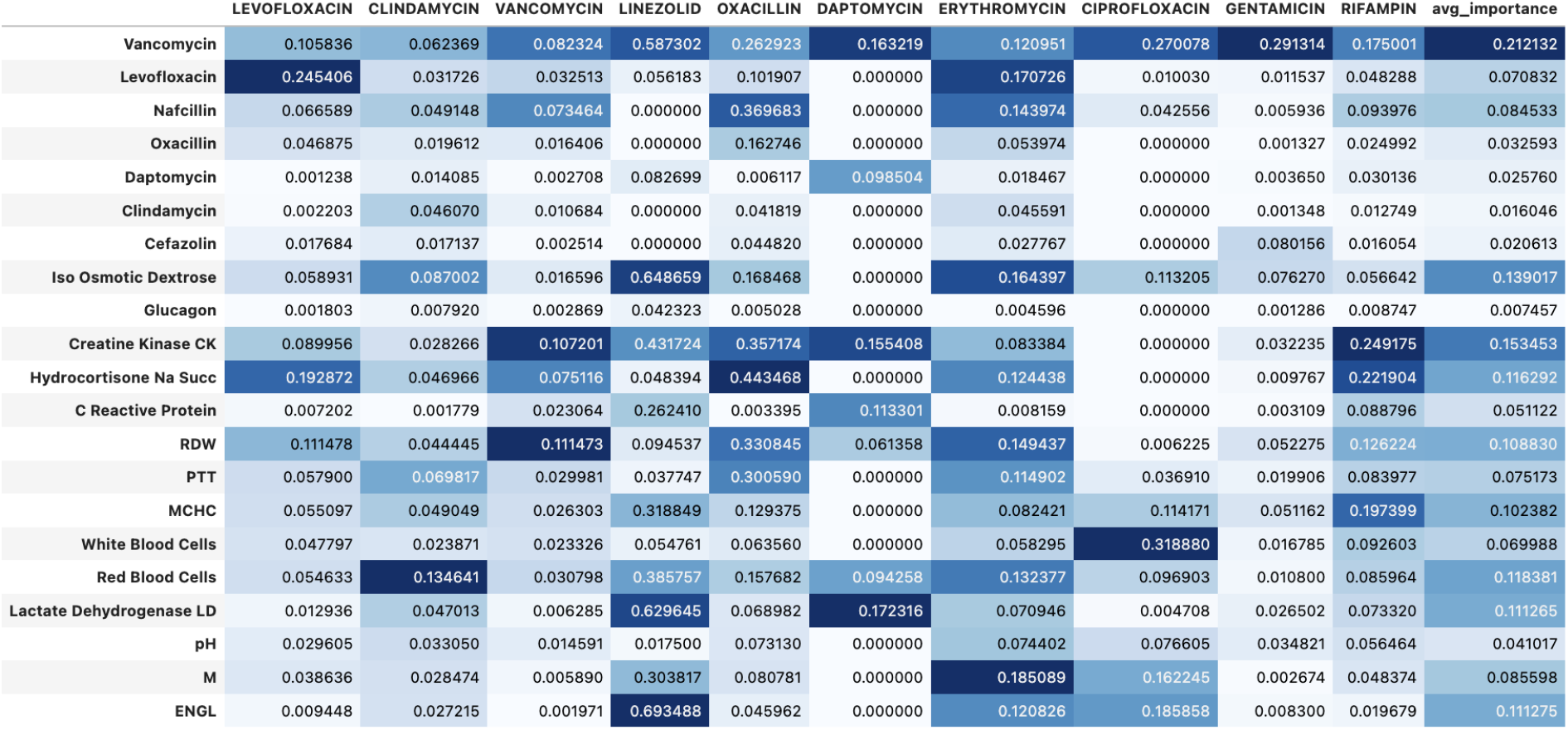
Variable Importances for each of the antibiotics found from Shapley Tree based variable importance calculations.

## 4. Discussion

In this study, we develop a novel method to extract raw clinical data and patient features from ICU admissions who are suspected to have a staph infection and show that we can apply data-driven methodologies to increase the efficiency and effectiveness of empiric antibiotic prescription. Our findings demonstrate that empiric antibiotic regimens have an efficiency rate of approximately 40 percent and specific regimens that are supposed to be tailored to address the susceptibility profile of the bacterial infection have an efficiency rate of 45 percent. The 9 antibiotic classifiers that we have constructed are able to utilize a patient’s entire electronic health record to predict whether an antibiotic will be active against the bacterial infection with an average accuracy of 85 percent. In all but 10 events (0.01 percent of the data) where an inappropriate antibiotic was prescribed, our model was able to identify at least one antibiotic that would have provided activity against the staph infection with an average of 3.5 antibiotics per inappropriate prescription. We have also identified that the prior antibiotic history of the patient, relevant blood test results, and demographic information such as age and language are among the most important features in the classification of antibiotic susceptibility. Moreover, we found that demographic information increases the accuracy of the model across all classifiers suggesting that race, language, and insurance are relevant factors that affect antibiotic susceptibility.

We believe that the implementation of an antibiotic susceptibility classifier for staph infections will prove useful for clinicians due to the identifiable phenotype of staph infections and the rising number of antibiotic-resistant strains. The process of treating a staph infection begins with identifying the relevant symptoms such as skin rashes, swelling of the skin, and red painful bumps. Then, the clinicians will order a microbiology test to identify the bacterial organism and a susceptibility test to guide therapy. Results from these tests are available within 76 hours. Although a quick recovery is anticipated with prompt treatment, there is a higher risk for severe problems the longer treatment is delayed - such as sepsis, pneumonia, endocarditis, and other bloodstream infections. For this reason, the treatment of staph infections usually begins with an empiric antibiotic prescription prior to the availability of lab tests. The doctor will perform an assessment of the patient’s health and the severity of the staph infection, and then, relying on clinical intuition and institution-wide antibiograms that track the susceptibility of isolated staph infections, they will most often prescribe Vancomycin, a first-line agent, or broad spectrum fluoroquinolones such as Ciprofloxacin or Levofloxacin. ^27,28,29,30^ However, the over-prescription of Vancomycin, one of the only drugs that can treat MRSA, has resulted in the development of Vancomycin-resistant strains of staph, and due to their overuse, many strains have similarly developed resistance to fluoroquinolones.^31^ Thus, our model will aid empiric therapy in order to prescribe more effective and specific antibiotics - increasing the quality of care and decreasing antibiotic resistance and staph-related mortality.

Numerous prior studies have demonstrated the need and the effectiveness of EHR-based machine learning models and clinical decision support systems to predict antibiotic resistance patterns, susceptibility, risk of infection, and conditions such as sepsis and mortality. For example, Kanjilal et al built an antibiotic stewardship algorithm to limit the number of inappropriate first-line antibiotics that were prescribed in favor of second-line prescriptions that were predicted to have the same coverage rate. Corbin et al developed machine-learning models to predict personalized antibiograms to aid in precise prescribing for the treatment of uncomplicated urinary tract infections, and Eickelberg et al identified patients with a low risk of bacterial infection to decrease the duration of empiric antibiotic therapy.^19,20^ Our study adds to the validity and the body of research surrounding EHR-based prediction models and supports the findings that it is possible to use electronic health records to aid in specific and effective empiric antibiotic therapy.

**Figure 11.**
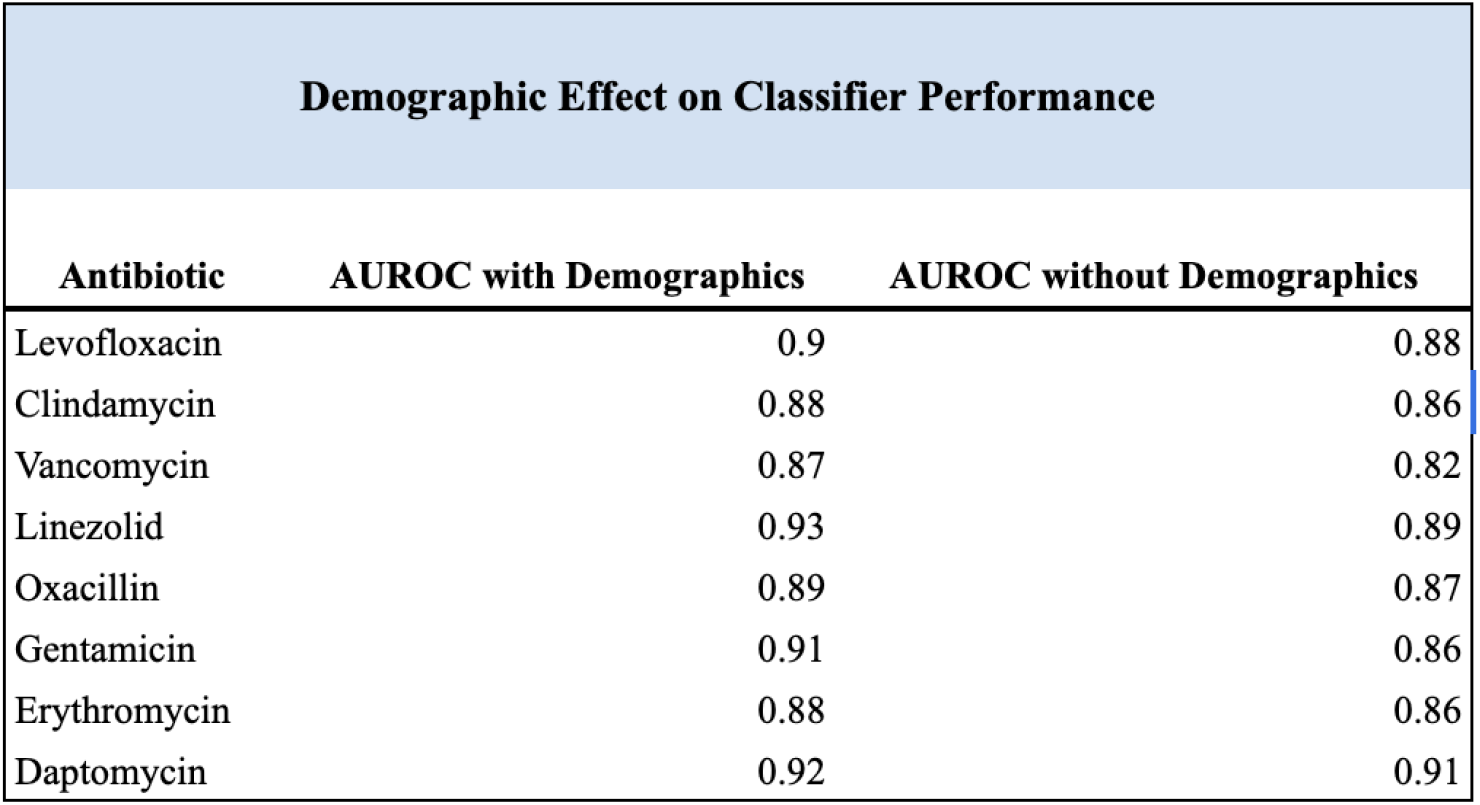
The results from the same antibiotic classifier including and withholding demographic information such as ethnicity, insurance, and language.

There are several limitations of this study. First, the data that was used was collected for clinical care at a single medical institution between the years 2001 and 2012 so it does not reflect current real-world antibiotic susceptibility patterns or clinician allocations. Moreover, a majority of the common and advised treatment regimes for staph infections such as nafcillin and cefazolin were not tested for susceptibility so we were not able to include these in our study.

Our future work will focus on the measures needed for the implementation of the antibiotic stewardship model in clinical practices and how to best integrate these clinical decision support systems into clinical workflows. Moreover, we will focus on generalizing these results to a broader patient population and further identify the discrepancies and trends with the various social determinants of health in our model in order to address health disparities and decrease the bias in our models. Moreover, to explore the longitudinal and temporal trends in the data we will investigate the use of deep learning models such as LSTM and GRU which are able to incorporate time-based information into the classification process.

## Data Availability

All data produced in the present study are available upon reasonable request to the authors.

https://physionet.org/content/mimiciii/1.4/

## Glossary

EHR: Electronic Health Record
MRSA: Methicillin-Resistant Staphylococcus Aureus
MIMIC−III: Medical Information Mart for Intensive Care III dataset

## Notes

### Competing Interest Statement

The authors have declared no competing interest.

### Funding Statement

This study did not receive any funding.

### Author Declarations

The MIMIC-III dataset was openly available and was initiated before the start of our study. The data is available at https://physionet.org/content/mimiciii/1.4/

## References

[1] Ventola, C Lee. “The antibiotic resistance crisis: part 1: causes and threats.” P T : a peer-reviewed journal for formulary management vol. 40,4 (2015): 277–83.

[2] Coculescu, Bogdan-Ioan. “Antimicrobial resistance induced by genetic changes.” Journal of medicine and life vol. 2,2 (2009): 114–23.

[3] Ziólkowski, Grzegorz et al. “Antibiotic Stewardship in Staphylococcus aureus Bloodstream Infection Treatment-Analysis Based on 29,747 Patients from One Hospital.” Antibiotics (Basel, Switzerland) vol. 9,6 338. 18 Jun. 2020.

[4] Bebell, Lisa M, and Anthony N Muiru. “Antibiotic use and emerging resistance: how can resource-limited countries turn the tide?.” Global heart vol. 9,3 (2014): 347–58. doi:10.1016/j.gheart.2014.08.009

[5] Tan, S.Y., Khan, R.A., Khalid, K.E. et al. Correlation between antibiotic consumption and the occurrence of multidrug-resistant organisms in a Malaysian tertiary hospital: a 3-year observational study. Sci Rep 12, 3106 (2022).

[6] Calderón-Parra, Jorge et al. “Inappropriate antibiotic use in the COVID-19 era: Factors associated with inappropriate prescribing and secondary complications. Analysis of the registry SEMI-COVID.” PloS one vol. 16,5 e0251340. 11 May. 2021, doi:10.1371/journal.pone.0251340

[7] “Antibiotic Resistance.” World Health Organization, World Health Organization, https://www.who.int/news-room/fact-sheets/detail/antibiotic-resistance.

[8] Pollack, Loria A, and Arjun Srinivasan. “Core elements of hospital antibiotic stewardship programs from the Centers for Disease Control and Prevention.” Clinical infectious diseases : an official publication of the Infectious Diseases Society of America vol. 59 Suppl 3,Suppl 3 (2014): S97–100. doi:10.1093/cid/ciu542

[9] Vincent, Jean-Louis et al. “International study of the prevalence and outcomes of infection in intensive care units.” JAMA vol. 302,21 (2009): 2323–9. doi:10.1001/jama.2009.1754

[10] Murphy, Marion et al. “Antibiotic prescribing in primary care, adherence to guidelines and unnecessary prescribing–an Irish perspective.” BMC family practice vol. 13 43. 28 May. 2012, doi:10.1186/1471-2296-13-43

[11] What Exactly is Antibiotic Resistance? (2022, October 5). Centers for Disease Control and Prevention. https://www.cdc.gov/drugresistance/about.html

[12] Algammal, Abdelazeem M et al. “Methicillin-Resistant Staphylococcus aureus (MRSA): One Health Perspective Approach to the Bacterium Epidemiology, Virulence Factors, Antibiotic-Resistance, and Zoonotic Impact.” Infection and drug resistance vol. 13 3255–3265. 22 Sep. 2020, doi:10.2147/IDR.S272733

[13] Okwu, Maureen U et al. “Methicillin-resistant Staphylococcus aureus (MRSA) and anti-MRSA activities of extracts of some medicinal plants: A brief review.” AIMS microbiology vol. 5,2 117–137. 15 Apr. 2019, doi:10.3934/microbiol.2019.2.117

[14] “An Estimated 1.2 Million People Died in 2019 From.” University of Oxford, https://www.ox.ac.uk/news/2022-01-20-estimated-12-million-people-died-2019-antibiotic-resistant-bacterial-infections.

[15] Durand, Claire et al. “Clinical Decision Support Systems for Antibiotic Prescribing: An Inventory of Current French Language Tools.” Antibiotics (Basel, Switzerland) vol. 11,3 384. 14 Mar. 2022, doi:10.3390/antibiotics11030384

[16] Davenport, Thomas, and Ravi Kalakota. “The potential for artificial intelligence in healthcare.” Future healthcare journal vol. 6,2 (2019): 94–98. doi:10.7861/futurehosp.6-2-94

[17] Goss, Foster R et al. “Improved antibiotic prescribing using indication-based clinical decision support in the emergency department.” Journal of the American College of Emergency Physicians open vol. 1,3 214–221. 13 Mar. 2020, doi:10.1002/emp2.12029

[18] Corbin, Conor K et al. “Personalized antibiograms for machine learning driven antibiotic selection.” Communications medicine vol. 2 38. 8 Apr. 2022, doi:10.1038/s43856-022-00094-8

[19] Eickelberg, Garrett et al. “Predictive modeling of bacterial infections and antibiotic therapy needs in critically ill adults.” Journal of biomedical informatics vol. 109 (2020): 103540. doi:10.1016/j.jbi.2020.103540

[20] Kanjilal, Sanjat et al. “A decision algorithm to promote outpatient antimicrobial stewardship for uncomplicated urinary tract infection.” Science translational medicine vol. 12,568 (2020): eaay5067. doi:10.1126/scitranslmed.aay5067

[21] Johnson, A., Pollard, T., Shen, L. et al. MIMIC-III, a freely accessible critical care database. Sci Data 3, 160035 (2016). https://doi.org/10.1038/sdata.2016.35

[22] Tong, Steven Y C et al. “Staphylococcus aureus infections: epidemiology, pathophysiology, clinical manifestations, and management.” Clinical microbiology reviews vol. 28,3 (2015): 603–61. doi:10.1128/CMR.00134-14

[23] Kwiecinski, Jakub M, and Alexander R Horswill. “Staphylococcus aureus bloodstream infections: pathogenesis and regulatory mechanisms.” Current opinion in microbiology vol. 53 (2020): 51–60. doi:10.1016/j.mib.2020.02.005

[24] Pedregosa, F. et al. Scikit-learn: machine learning in Python. J. Machine Learning Res. 12, 2825–2830 (2011).

[25] Ke, G. et al. Lightgbm: a highly efficient gradient boosting decision tree. Adv. Neural Inform. Processing Syst. 30, 3146–3154 (2017).

[26] Li, Xiaoxiao et al. “Efficient Shapley Explanation For Features Importance Estimation Under Uncertainty.” Medical image computing and computer-assisted intervention : MICCAI … International Conference on Medical Image Computing and Computer-Assisted Intervention vol. 12261 (2020): 792–801. doi:10.1007/978-3-030-59710-877

[27] Rasmussen, Rasmus V et al. “Future challenges and treatment of Staphylococcus aureus bacteremia with emphasis on MRSA.” Future microbiology vol. 6,1 (2011): 43–56. doi:10.2217/fmb.10.155

[28] American Academy of Pediatrics. Staphylococcal Infections. (https://www.healthychildren.org-issues-Infections.aspx) Accessed 11/25/2022.

[29] Centers for Disease Control and Prevention. Methicillin-resistant Staphylococcus aureus (MRSA). (https://www.cdc.gov/mrsa/index.html) Accessed 11/25/2022.

[30] Lowy FD. Lowy F.D. Lowy, Franklin D.Staphylococcal Infection. In: Jameson J, Fauci AS, Kasper DL, Hauser SL, Longo DL,Loscalzo J. Jameson J, Fauci A.S., Kasper D.L., Hauser S.L., Longo D.L., Loscalzo J Eds. J. Larry Jameson, et al.eds. Harrison′sPrincipleso f InternalMedicine, 20eNewYork, NY : McGraw–Hill.Accessed11/25/2022.

[31] Cong, Yanguang et al. “Vancomycin resistant Staphylococcus aureus infections: A review of case updating and clinical features.” Journal of advanced research vol. 21 169–176. 12 Oct. 2019, doi:10.1016/j.jare.2019.10.005

